# Multidimensional apathy: A simple and inclusive clinical marker of youth mental health—A longitudinal study

**DOI:** 10.1101/2025.03.10.25323681

**Authors:** Giulia Lafond-Brina, Anne Bonnefond

## Abstract

**Background:** Early identification and treatment of young individuals at risk for psychiatric disorders are essential to reducing the burden of mental health disorders, which are among the leading causes of disability worldwide. This study aims to determine whether different phenotypes of multidimensional apathy could be potential predictors to the transition to distinct psychiatric disorders in youth and whether they may serve as clinical markers.

**Methods:** In a longitudinal study, we followed 470 young adults over a period of 4.5 years. They completed online surveys, providing self-reported diagnoses of psychiatric disorders along with socio-demographic, medical, educational, and functional characteristics. Multidimensional apathy, self-esteem, depressive symptoms, and motivation were assessed using validated scales. Binary logistic regression analyses were conducted to identify predictors of psychotic or depressive disorders at the 4.5-year follow-up, while a Chi-squared test examined the stability of multidimensional apathy over time.

**Results:** The presence of a self-diagnosed psychotic disorder in 2024 was predicted in 2020 by a low self-esteem (β=−0.19;p<3.9×10−8), consumption of legal psychoactive drugs (β=1.13;p<0.002), an absence of anxiety (β=−2.22;p<0.003), higher emotional apathy (β=1.83;p<0.006), lack of leisure (β=−1.774;p<0.02), antecedents of psychiatric disorders in first-degree relatives (β=1.505;p<0.03), and consumption of illicit substances other than cannabis (β=1.726;p<0.03), with an overall accuracy of 76.81%. The presence of a depressive disorder was predicted by subclinical depressive symptoms (β=5.672;p<2.3×10−6), being a junior in university (β=3.681;p<0.003), presence of non-psychiatric disorders (β=1.899;p<0.03), higher executive apathy (β=2.229;p<0.03), consumption of alcohol (β=2.128;p<0.06), and lack of independence (β=−1.867;p<0.07), with an overall accuracy of 91.47%. Participants presented a temporal stability for emotional and executive apathy between 2020 and 2024 (p<2.2×10−16).

**Conclusions:** Our results show for the time emotional and executive apathy as predictors of the transition to psychosis and depression, respectively, at 4.5 years, suggesting multidimensional apathy as an easy-to-implement and inclusive candidate for clinical markers of youth mental health.

## 1. Introduction

Youth mental health, as recently emphasized by The Lancet Psychiatry Commission, must be recognized as a public health priority ^1^. Mental illness affects approximately 30% of individuals aged 18 to 20, with the period from puberty to the mid-to-late twenties representing a critical time of vulnerability for the onset of psychiatric disorders ^2^. Notably, 63–75% of mental disorders emerge before the age of 25, highlighting the urgent need for early detection and intervention strategies. Indeed, early interventions have been shown to reduce the incidence of mental disorders by improving long-term symptomatic and functional outcomes, lowering suicide rates following diagnosis, and causing a delayed or less disabling transition to psychosis ^1,3,4^. Consequently, the need for appropriate care arises well before a formal diagnosis, which typically occurs between ages 25–27 for schizophrenia and 30–35 for mood disorders ^5^.

To advance research regarding the prediction of youth mental health disorders, identifying new and inclusive clinical markers—more cost-effective than genetic or functional biomarkers and more readily applicable in clinical practice—is crucial. Indeed, sudden psychiatric disorders are often preceded by early symptoms, which may serve as personalized clinical biomarkers ^6^. Clinically, these early symptoms appear during youth and manifest as (micro)phenotypes, that may fluctuate in severity and specificity throughout the transition to a full-blown disorder. However, they do not necessarily follow distinct trajectories leading to specific diagnoses such as schizophrenia or depression ^7^.Conducting longitudinal studies with transdiagnostic models in emerging adulthood can therefore facilitate the understanding of these early symptoms and their trajectories.

Moreover, a longitudinal perspective allows the exploration of mechanisms that underpin mental ill health. This framework places people along a multidimensional continuum of health to illness, capturing elements of risk, onset, course, and prognosis. The current models of psychosis and mood disorder development suggest that they result from the complex interplay of genetic and environmental factors, which can either increase (risk factors) or decrease (protective factors) the likelihood of developing psychiatric disorders ^8^. Among these, “non-purely genetic factors” encompass numerous influences, including socio-demographic variables, parental and perinatal antecedents, later-life factors and personal conditions ^9^. Concerning socio-demographic factors, it is important to note that in low-resource and middle-resource countries, mental healthcare and prevention systems for youth are typically absent or limited because global research propositions are often highly expensive ^1^, even if poverty is a well-known risk factor of transition to psychiatric disorders ^9^.

Despite the numerous putative risk factors, the literature favors more non-specific genetic and “non-purely genetic” factors that transcend each categorical diagnosis. For example, several potential risk factors have been identified in genetics and imaging studies, but mostly on the psychiatric spectrum, independently of the diagnostic disorder ^10–12^. In clinical biomarkers, the anhedonia trait appears to be a common early risk factor of both psychotic disorders ^8,9^, and depressive disorders ^13,14^. Previous studies have also found that apathy is an early common risk factor for the development of psychotic and mood disorders ^15–17^; being a transdiagnostic negative symptom, apathy is a considerably disabling symptom in psychiatry ^18^.

However, we believe that the exploration of multidimensional apathy, which is almost non-existent in psychiatry, could improve clinical guidance during emerging adulthood, by predicting specific psychiatric disorders. It is a timely subject since 27% of youth at high risk of psychosis will transition to psychosis after four years, while the remaining 73% will develop other mental disorders, especially in the mood-spectrum disorders ^19^. A multidimensional model of apathy has demonstrated its clinical relevance in neurology, by distinguishing three phenotypes—emotional, executive, and initiative—with specific symptoms ^20^ underlying distinct networks ^21,22^. Patients with emotional apathy suffer from an emotional blunting and indifference, while those with executive apathy suffer from a difficulty to plan actions and those with an initiative apathy have difficulty in initiating thoughts, actions, or emotions. We recently showed that the three phenotypes of apathy exist isolated in healthy young adults, with the same disabling consequences as the apathy symptom in disorders; moreover, each was associated with specific predictors and specific cognitive mechanisms in these subjects ^23–25^.

Therefore, the current aim is to test if different phenotypes of multidimensional apathy could predict the transition to distinct psychiatric disorders in youth and if these phenotypes could even be clinical markers. Indeed, more than a risk factor, a clinical marker must be a stable, non-contextual trait that reveals underlying mechanisms of a disorder and informs the development of more targeted treatments ^26,27^. Moreover, a clinical marker has to be inexpensive, simple, and possibly digital to compete with genetic and imaging markers and overcome the current global limitation of healthcare and disease prevention being accessible to only high and middleclass consumers ^1^. Therefore, the clinical importance of investigating multidimensional apathy is threefold. First, the three phenotypes could potentially be used to improve and refine the prediction of transition to psychosis and depression in the general population, especially among people at risk of developing psychiatric disorders. Second, these risk factors may be clinical markers that can be potentially modifiable by preventive interventions. Third, these inclusive and inexpensive clinical factors are easy to measure over time using questionnaires and could be easily utilized for promoting worldwide mental health.

## 2. Method

### 2.1. Participants

In total, 624 young adults responded to the online survey about the “evolution of personality in young adults” conducted in June 2024. Each respondent had previously participated in a 2020 survey focusing on “personality in university students”, conducted on the entire student population of a French University, and had agreed to be contacted again by email at a later date (the results of the first survey are published in Lafond-Brina et al., 2022). After eliminating those who did not answer all the questions, 470 participants constituted the total sample.

To meet our objective, all participants of this sample who, in the initial survey, reported suffering from a psychiatric disorder and/or presented with moderate or severe depression (score superior to 13 on the Beck Depression Inventory II [BDI-II] ^28–30^ were excluded. Overall, 341 subjects constituted the sample of interest.

Figure 1 represents the flowchart of the participants’ inclusion, while Table 1 summarizes the demographic and neuropsychological characteristics of the total sample in 2020 and 2024 and the sample of interest in 2024.

**Figure 1.**
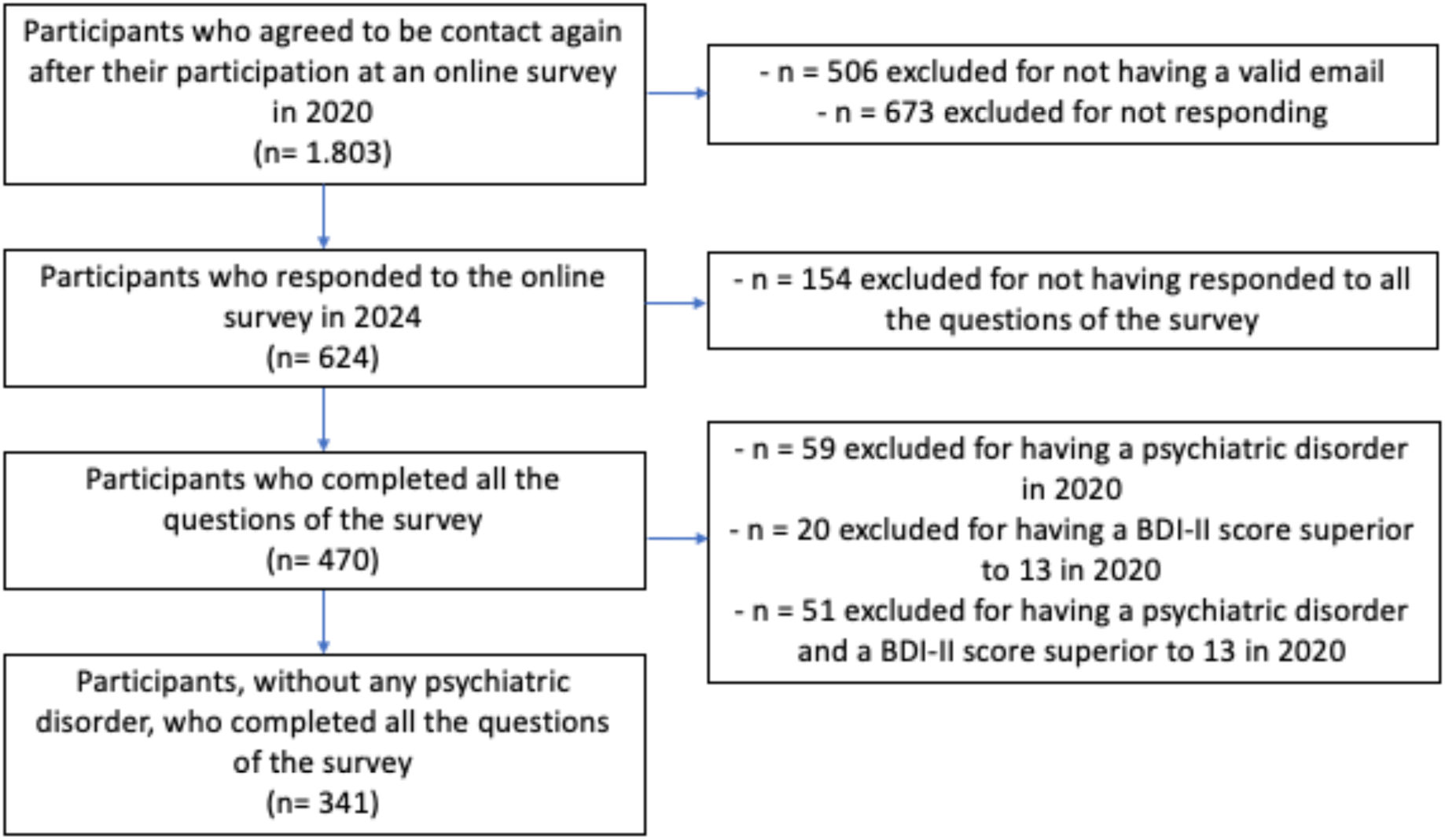
Flowchart of participants’ inclusion and selection process

**Table 1:** Demographic and neuropsychological characteristics of the 470 subjects in 2020 and 2024. Values are given as the mean (standard deviation), the Dimensional Apathy Scale [DAS], the Beck Depression Inventory II [BDI-II], the Rosenberg Self-Esteem Scale [RSE], the Temporal Experience of Pleasure Scale [TEPS].

|  | 2020 February<br>n = 470 | 2024 June<br>n =470 | 2024 June<br>n = 341 |
| --- | --- | --- | --- |
| Age (years) | 20.3 (2.22) | 24.9 (2.22) | 25.3 (2.14) |
| Sex (% of males) | 26.6 | 26.6 | 29.7 |
| Level of education | Freshman: 29.1%<br>Junior: 41.9%<br>Senior: 28.9% | Freshman: 5.8%<br>Junior: 25.6%<br>Senior: 68.7% | Freshman: 5.6%<br>Junior: 27.0%<br>Senior: 67.3% |
| DAS executive | 11.1 (5.09) | 10.7 (5.11) | 9.5 (4.73) |
| DAS emotional | 7.79 (4.35) | 7.06 (4.16) | 7.19 (4.23) |
| DAS initiative | 10.10 (3.94) | 9.93 (3.73) | 9.71 (3.64) |
| BDI-II | 8.60 (6.77) | 8.16 (6.32) | 6.53 (5.16) |
| RSE | 27.2 (6.27) | 28.2 (6.23) | 29.6 (5.82) |
| TEPS Anticipatory pleasure | 44.8 (7.05) | 44.9 (7.03) | 45.4 (6.82) |
| TEPS Consummatory pleasure | 37.1 (5.89) | 38.4 (5.78) | 38.6 (5.79) |

All participants gave electronic informed consent and were not paid for their participation. The study was approved by the University review board (UNISTRA/CER/2020-13) and the French data protection authority (CNIL).

### 2.2. Variables of reference

The presence of a psychotic disorder in 2024 was deduced from the answer to the question “Since the last survey in 2020, do you suffer, or have you suffered from a psychiatric disorder such as schizophrenia or bipolarity?”. Two answers were proposed: “yes” and “no”.

The presence of a depressive disorder in 2024 was deduced from the BDI-II score. According to the creators of the scale and some articles with the same population, the cut-off of 13 on the BDI-II allow to distinguish two groups: (1) presence of a moderate to severe depressive disorder if the BDI-II score in 2024 was superior to 13 and (2) absence of a depressive disorder if the BDI-II score in 2024 was inferior or equal to 13^28–30^.

### 2.3. Regressors of interest

The regressors of interest, which can be grouped into five categories, were:

1. sociodemographic characteristics: gender, age, and level of education
2. medical history: presence of non-psychiatric disorders (such as physical or sensory disability, thyroid disorder, neurological disease), antecedents of psychiatric disorder in first-degree relatives, consumption of legal psychoactive drugs (such as anxiolytics and sleeping pills), consumption of alcohol, consumption of cannabis, consumption of substances other than alcohol or cannabis
3. education: current field studied at the university, number of repeated grades, number of skipped grades, and presence of professional- or university-related anxiety
4. general functioning: type of accommodation, number of initiated contacts, presence of social isolation, presence of leisure, number of outings per month, daily fatigue, curiosity, independence, pleasure in daily life, realization of chores, realization of the grocery shopping, realization of the home cooking, and number of sedentary hours per day
5. psychopathology: scores of four validated scale assessed multidimensional apathy, self-esteem, depressive symptoms, and motivation: DAS executive score, DAS emotional score, DAS initiative score, RSE score, BDI score, TEPS anticipatory score and TEPS consummatory score respectively (see Table 2)

**Table 2:** Description and interpretation of the four validated scales that assessed psychopathology.

| Questionnaires | Description | Interpretation |
| --- | --- | --- |
| The Dimensional Apathy Scale (DAS) <sup>31</sup> | A 24-item scale assessing the severity of the three phenotypes of apathy (executive, emotional, and initiative apathy). Items are scored on a 4-point Likert scale based on the frequency of occurrence of the apathetic symptoms in the previous month. | 3 scores are obtained, one for each phenotype of apathy (based on 8 items each). A high score (maximum 24) indicates a severe phenotype of apathy. |
| The Beck Depression Inventory II (BDI-II) (Beck et al., 1996) | A 13-item scale assessing the severity of depressive symptoms. Each item is scored from 0 to 3. | A high global score (maximum 39) indicates severe depression. A score >13 indicates moderate or severe depression. |
| The Rosenberg Self-Esteem Scale (RSE) <sup>32</sup> | A 10-item scale assessing self-esteem. Each item is scored from 1 to 4. | A low global score (minimum 10) indicates severe self-esteem disorder. |
| The Temporal Experience of Pleasure Scale (TEPS) <sup>33</sup> | An 18-item scale assessing anticipatory (10 items) and consummatory (8 items) pleasure. Each item is scored from 1 to 6. | A low score on the anticipatory pleasure subscale indicates difficulty in actively seeking or anticipating enjoyable activities (score range: 10–60). A low score on the consummatory pleasure subscale indicates difficulty in experiencing immediate pleasure from completing enjoyable activities (score range: 8–48). |

In Supplementary Data 1, a description of the qualitative regressors of interest of the five categories is detailed.

### 2.4. Statistical analysis

The percentage of subjects in the sample that had a psychotic disorder, a depressive disorder, or both in 2024 were calculated. Moreover, binary logistic regression analyses were used to analyze the predictors of the presence of a psychotic or depressive disorders. Using a mixed stepwise selection, we aimed to determine which variables in the survey most influenced the variable of reference due to the Akaike Information Criterion (AIC) —smaller AIC values indicate better the model fit, using the fewest possible variables. Furthermore, R-square values (the percentage variation of the reference explained by the regressor of interest) and the variance inflation factor (VIF), which measures the amount of multicollinearity in the regressors of interests, were calculated—higher VIF indicates higher possibility that multicollinearity exists. In binary logistic regression, VIF values above 2.5 may be a cause for concern ^34^. A confusion matrix was then proposed to determine the accuracy, the Cohen’s kappa, the sensitivity, the specificity, and the positive and negative predictive values of the models.

A complementary analysis was proposed to test the stability of the multidimensional apathy trait. Since some intra-subjects’ variations in questionnaires can be clinically insignificant, a Chi-squared test was used to determine whether the proportion of subjects remaining in the same category (subclinical apathy, or lack of apathy) in 2024 than in 2020 was significantly different from the expectations.

## 3. Results

### 3.1. Predictors of psychiatric disorders

#### 3.1.1. Characterization of the sample with a psychotic disorder, a depressive disorder, or both in 2024

Overall, 74 subjects developed a psychotic disorder between 2020 and 2024, while 42 subjects developed a moderate to severe depressive disorder between 2020 and 2024. Only 25 developed both a psychotic disorder and a moderate to severe depressive disorder. Thus, only 33.78% of participants with a psychotic disorder also presented a concomitant depressive disorder.

#### 3.1.2. Predictors of psychotic disorders

In a binary logistic regression model with the presence/absence of a psychotic disorder as the reference, RSE score (β=−0.193), consumption of legal psychoactive drugs (β=1.127), presence of professional- or university-related anxiety (β=−2.222), DAS emotional score (β=1.828), presence of leisure (β=−1.774), antecedents of psychiatric disorder in first-degree relatives (β=1.505), and consumption of substances other than alcohol or cannabis (β=1.726) explained 21.8% of the variance in the presence/absence of a psychotic disorder (X^2^(6)=45.3;p<.001;AIC=318.75; VIF=[1.03;1.58]). Table 3 depicts the statistical characteristics of the predictors. The confusion matrix analysis revealed an overall accuracy of 76.81% (95%CI: 72.72%– 80.55%), with a highly significant deviation from the no-information rate (p=8.547×10−7). The Cohen’s kappa was 0.44, and sensitivity was high at 89.46%, while specificity was lower at 51.59%. The positive predictive value was 78.65%, and the negative predictive value was 71.05%.

**Table 3:**
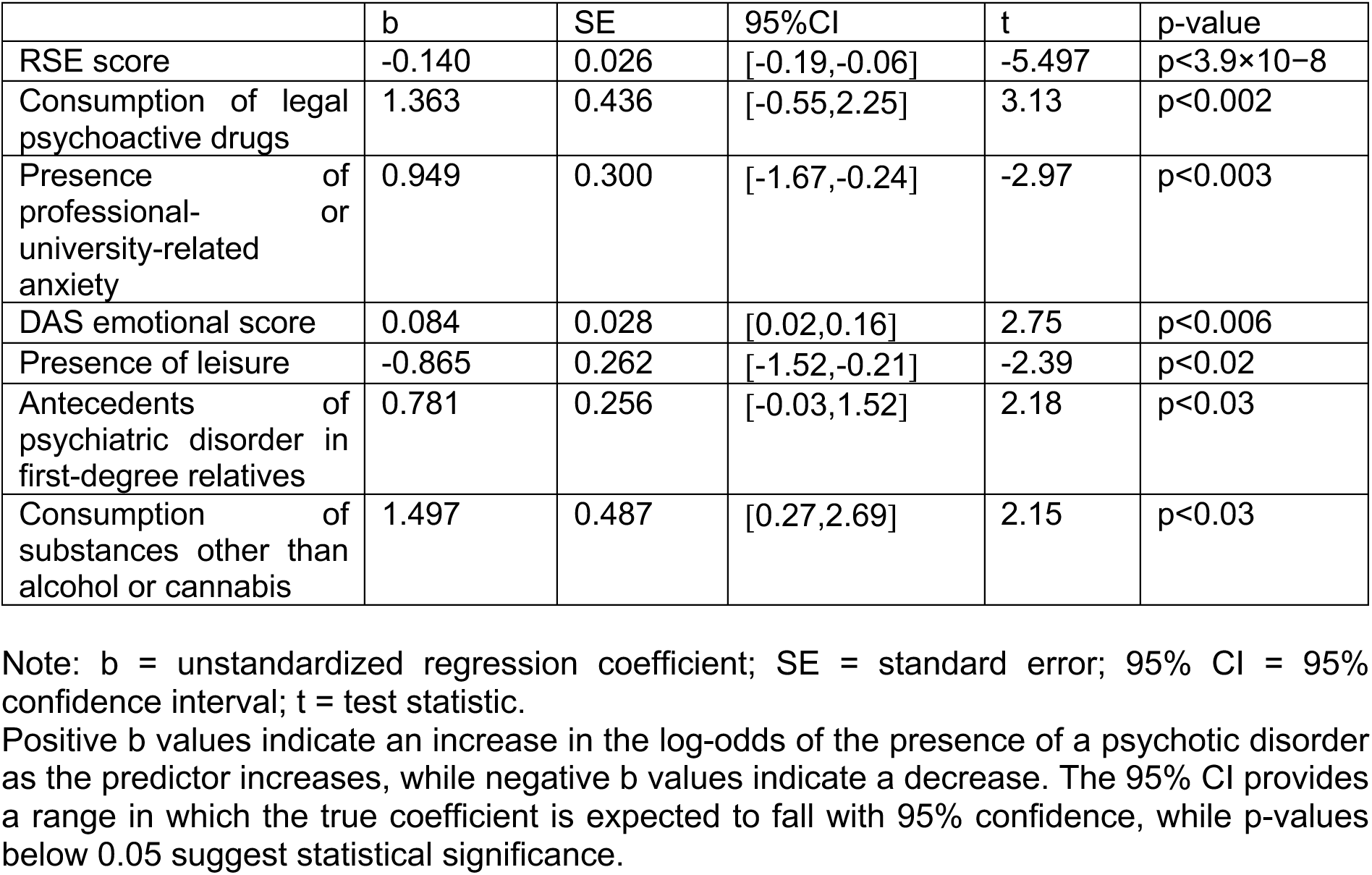
Predictors in 2020 of a psychotic disorder in 2024.

#### 3.1.3. Predictors of depressive disorders

In a binary logistic regression model with the presence/absence of a depressive disorder as the reference, subclinical BDI score (β=5.672), level of education (being junior in university; β=3.681), presence of non-psychiatric disorders (β=1.899), DAS executive score (β=2.229), consumption of alcohol (β=2.128), and independence (β=−1.867) explained 53.1% of the variance in the presence/absence of a depressive disorder (X^2^(5)=66.9;p<.001;AIC=205.06; VIF=[1.00;1.15]). Table 4 depicts the statistical characteristics of the predictors. The confusion matrix analysis revealed an overall accuracy of 91.47% (95%CI: 87.98%–94.21%), with a significant deviation from the no-information rate (p<0.003). The Cohen’s kappa was 0.52, and sensitivity was high at 97.99%, while specificity was lower at 45.24%. The positive predictive value was 92.70%, and the negative predictive value was 76.00%.

**Table 4:** Predictors in 2020 of a depressive disorder in 2024.

|  | b | SE | 95%CI | t | p-value |
| --- | --- | --- | --- | --- | --- |
| Subclinical BDI score | 0.240 | 0.051 | [0.15,0.35] | 4.73 | $p<2.25\times10^{-6}$ |
| Being junior in university | 1.607 | 0.535 | [0.30,2.71] | 2.99 | $p<0.003$ |
| Presence of non-psychiatric disorders | 1.335 | 0.622 | [0.12,2.55] | 2.15 | $p<0.03$ |
| DAS executive score | 1.147 | 0.547 | [-0.14,2.23] | 2.10 | $p<0.03$ |
| Consumption of alcohol | 0.136 | 0.073 | [-0.05,0.29] | 1.87 | $p<0.06$ |
| Independence | -0.841 | 0.480 | [-1.49,0.11] | -1.75 | $p<0.07$ |

Note: b = unstandardized regression coefficient; SE = standard error; 95% CI = 95% confidence interval; t = test statistic.
Positive b values indicate an increase in the log-odds of the presence of a depressed disorder as the predictor increases, while negative b values indicate a decrease. The 95% CI provides a range in which the true coefficient is expected to fall with 95% confidence, while p-values below 0.05 suggest statistical significance.

#### 3.1.4. Predictors of a psychotic disorder with a concomitant depressive disorder

In a binary logistic regression model with the presence/absence of both a psychotic and a depressive disorder as the reference, dynamism (β=−3.728), presence of non-psychiatric disorders (β=3.904), pleasure in daily life (β=−6.799), age (β=−5.508), absence of realization of home cooking (β=10.405), and TEPS consummatory score (β=−3.569) explained 42.8% of the variance in the presence/absence of both a psychotic and a depressive disorder as the reference (X^2^(5)=40.2;p<.001;AIC=154.12; VIF=[1.08;1.50]). Table 5 displays the statistical characteristic of the predictors. The confusion matrix analysis revealed an overall accuracy of 95.59% (95%CI: 92.83%– 97.51%), with a significant deviation from the no-information rate (p<0.009). The Cohen’s kappa was 0.59, and sensitivity was high at 99.37%, while specificity was lower at 48.00%. The positive predictive value was 96.01%, and the negative predictive value was 85.71%.

**Table 5:** Predictors in 2020 of a co-occurrence of psychotic and depressive disorders in 2024.

|  | b | SE | 95%CI | t | p-value |
| --- | --- | --- | --- | --- | --- |
| Dynamism | -1.286 | 0.712 | [-2.74,0.09] | -1.81 | $p<1.59\times 10^{-4}$ |
| Presence of non-psychiatric disorders | 2.259 | 0.849 | [0.62,4.00] | 2.66 | $p<5.93\times 10^{-4}$ |
| Pleasure in daily life | -1.782 | 0.632 | [-3.11,-0.59] | -2.82 | $p<8.32\times 10^{-4}$ |
| Age | -0.396 | 0.176 | [-0.61,-0.08] | -2.24 | $p<0.02$ |
| Absence of realization of home cooking | 3.313 | 1.179 | [1.17,5.86] | 2.81 | $p<0.03$ |
| TEPS consummatory score | -0.095 | 0.055 | [-0.01,0.21] | 1.74 | $p<0.04$ |

Note: b = unstandardized regression coefficient; SE = standard error; 95% CI = 95% confidence interval; t = test statistic.
Positive b values indicate an increase in the log-odds of the presence of a co-occurrence of psychotic and depressed disorders as the predictor increases, while negative b values indicate a decrease. The 95% CI provides a range in which the true coefficient is expected to fall with 95% confidence, while p-values below 0.05 suggest statistical significance.

### 3.2. Complementary analyses

#### 3.2.1. Longitudinal intra-subject evolution of apathy categorization (apathy vs no apathy)

The Chi-squared test for the given probabilities yielded a highly significant result for executive categorization (X^2^=162.08, df=1, p<2.2×10−16), indicating a significant tendency of subjects to remain in the same DAS executive category between 2020 and 2024.

The Chi-squared test for the given probabilities yielded a highly significant result for emotional categorization (X^2^=243.07, df=1, p<2.2×10−16), indicating a significant tendency of subjects to remain in the same DAS emotional category between 2020 and 2024.

The Chi-squared test for the given probabilities yielded a highly significant result for initiative categorization (X^2^=196.63, df=1, p<2.2×10−16), indicating a significant tendency of subjects to remain in the same DAS initiative category between 2020 and 2024.

## 4. Discussion

This longitudinal study, which followed a cohort of 470 young adults followed for four years, revealed for the first time that apathy, in a multidimensional conception, is an early predictor of interest for the specific transition to psychosis or depressive disorders in emerging adulthood. By highlighting that the emotional and executive phenotypes of apathy are predictive of the transition to psychosis and depression, respectively, at 4.5 years, our results have significant implications on the issue of youth mental health. Indeed, if clinical predictors of psychiatric disorders in youth have already been identified (predictors that were also found in the present study), none, to date, has been able to specifically predict the transition to a psychotic or depressive disorder and to meet all the criteria of a good clinical biomarker.

### Multidimensional apathy, as a specific predictor of psychiatric disorders, could offer new targeted therapeutic strategies

Indeed, our results show that emotional apathy can significantly impact the likelihood of developing a psychotic disorder within 4.5 years. The presence of a psychotic disorder, such as schizophrenia or bipolar disorder, was based on self-reported diagnoses. Among the predictors highlighted, emotional apathy and the absence of anxiety were the strongest (standardized coefficients β of 1.83 and −2.222, respectively). Notably, as shown in the initial survey of the same population in 2020, these two predictors are strongly correlated ^25^. Indeed, in young adults, lower anxiety, when accompanied by indifference toward this anxiety, was associated with greater severity of emotional apathy. Consequently, measures of emotional apathy and lack of anxiety may represent the same clinical difficulty and could therefore be underpinned by similar cognitive processes ^24^. Similarly, the results of the present study reveal that executive apathy significantly influences the likelihood of developing a depressive disorder within 4.5 years. The presence of a depressive disorder was based on self-reported responses to the BDI scale. Using the odds ratio of its b coefficient (1.147), our results show that individuals with higher levels of executive apathy are approximately 3.15 times more likely to experience a depressive disorder within 4.5 years than individuals without executive apathy.

So far, apathy—when defined as a reduction in goal-directed behavior—has been present since the earliest descriptions of untreated youth at risk of schizophrenia or depression ^35–37^ and is recognized as one of the most prevalent symptoms in psychiatric disorders ^18,38^. However, this classical evaluation of apathy has not been able to specifically predict psychiatric disorders in cross-sectional studies of high-risk youth ^16,17^. Contrarily, our results suggest that a finer assessment of the specific domain impacted by apathy—whether cognitive, emotional, or related to auto-activation ^39^—makes such prediction possible. Although the multidimensional conception of apathy is rarely applied in psychiatry to our knowledge, it is well established that a continuum of abnormalities in goal-oriented behaviors underlies different phenotypes of apathy, each associated with distinct neural and cognitive alterations ^20,21,24^. More specifically, orbitofrontal and subgenual cingulate dysfunction contribute to emotional apathy, which, on a more functional level, is associated with a “disliking” impairment, i.e. an indifference to negative information. Executive apathy, however, is associated with ventrolateral prefrontal cortex and posterior parietal dysfunction and an impairment of proactive control mode, corresponding to an anticipatory engagement of cognitive control processes. This knowledge makes this predictor an excellent potential clinical marker since the use of targeted therapies, a major point for improving outcomes in at-risk youth, becomes a real possibility. Encouraging results are already supporting this direction. For example, a recent study showed that rTMS stimulation of the orbitofrontal cortex could represent a promising therapeutic alternative in young adults with schizophrenia, by revealing a significant reduction of psychotic and negative symptoms in patients with drug-naïve, first-episode schizophrenia ^40^. Concerning depressive disorders, the first, tenuous argument arose from an fMRI study which showed that a digital intervention specifically designed to enhance cognitive control processes was effective in reducing depressive and apathetic symptoms, as well as in normalizing activity of the cognitive control network^41^. In contrast, a meta-analysis of cognitive remediation targeting a broad range of executive functions in individuals with depression found no significant reduction in the severity of depressive symptoms ^42^.

### Emotional and executive apathy as promising clinical markers

In addition to promising treatment strategies, emotional and executive phenotypes of apathy, unlike the other predictors identified, meet all the criteria of a good clinical biomarker. Indeed, starting with familial and personal antecedents, the most frequent predictor of the transition to a psychiatric disorder identified in the literature, our results showed that antecedents in first-degree relatives predict psychotic disorders, whereas personal medical antecedents predict depressive disorders. However, no study, to our knowledge, has shown that these antecedents were able to predict the transition towards a specific psychiatric disorder ^43–47^. Moreover, by showing a strong association between antecedents in first-degree relatives and both psychosis and resistant depression, genetics studies have confirmed the non-specificity of an antecedent in first-degree relatives ^10,11,48^. However, this phenotype of resistant depression usually emerges in the thirties and could therefore have been missed in our sample. Consequently, although a history of psychiatric disorders in first-degree relatives is a potential predictor of both psychosis and resistant depression, our findings could only reflect its association with psychosis. Furthermore, substances intake, specifically the consumption of alcohol and of both psychoactive medication and illicit drugs, have been identified as predictors of depressive and psychotic disorders, respectively. However, the risks that they may be more predictive of comorbidities should be emphasized. Indeed, substance abuse disorders and sleep disorders are frequent comorbidities of both psychotic and depressive disorders ^49–57^. Although no direct sleep measures were included in the survey, the use of legal psychoactive drugs could serve as an indirect indicator of sleep disorders, as these substances are primarily used for sleep management among emerging adults in France ^58,59^. Moreover, the survey does not provide information on the type and frequency of drug use, which prevents exploration of the potential dose-response relationship between the consumption of these substances and the risk of developing a psychotic disorder.

Finally, some psychological factors measured by questionnaires, such as a lack of self-esteem and subclinical depressive symptoms, have also been identified as predictors of psychotic and depressive disorders, respectively. However, the longitudinal follow-up at four years showed that the emotional and executive phenotypes of apathy were stable in time, whereas self-esteem and subclinical depressive symptoms were not. In addition to this temporal stability, our previous results regarding young adults showed that emotional and executive apathy were stable before and during the COVID-19pandemic, revealing a contextual stability, unlike self-esteem and depressive symptoms ^25^. Multidimensional apathy also seems to be an inclusive marker, showing cultural stability across young and older adults from various countries ^60,61^. On the contrary, the non-stability over time, contexts and cultures for self-esteem and depressive symptoms has been confirmed by several studies ^62–65^. Besides its stability, multidimensional apathy can also be easily integrated into clinical practices. Indeed, validated scales for assessing its intensity, like DAS, are available for use with youth from diverse countries ^25,60,66^. Furthermore, this symptom can also be objectively measured using (digital) behavioral markers ^67–69^. Behavioral measures offer the added advantage of requiring less metacognitive effort, which is particularly relevant since youth at psychiatric risk often exhibit reduced self-awareness of their symptoms compared to their healthy peers ^70^.

Our findings, based on self-reported diagnoses, need to be confirmed by a complementary longitudinal study using clinical assessments conducted by psychiatrists to provide a more robust understanding of the evolution of these young adults and the diagnostic trajectories within depressive and psychotic disorders.

In conclusion, identifying young persons at very early stages of psychiatric diseases, such as schizophrenia or mood disorders, is crucial to mitigate the growing worldwide impact of mental health disorders, one of the most common worldwide causes of disability. Our longitudinal results suggest that multidimensional apathy is a simple, easy-to-implement and inclusive candidate for clinical markers, which will help advance youth mental health prediction and personalize the strategies to target for improving the efficacy of preventive interventions. By stipulating “candidate,” we anticipate this topic will receive active consideration by the field, because, to date, not enough data are available to fully evaluate its status as a specific clinical marker to identify youth at risk of psychotic or depressive disorders.

## Data Availability

All data produced in the present study are available upon reasonable request to the authors.

## Acknowledgements

G. L-B and A. B designed the study. G. L-B realized the acquisition of the data, did the statistical analysis, and wrote the first draft of the manuscript. Both authors were involved in the writing of this draft.

